# Testing trajectories for GetCheckedOnline in British Columbia: Implications for equity, stewardship and sustainability of digital STI testing services

**DOI:** 10.64898/2026.04.12.26350729

**Authors:** Ihoghosa Iyamu, Devon Haag, Sofia Bartlett, Catherine Worthington, Daniel Grace, Mark Gilbert

**Affiliations:** School of Population and Public Health, The University of British Columbia, Vancouver, BC, Canada; BC Centre for Disease Control, Vancouver, British Columbia, Canada; School of Public Health and Social Policy, University of Victoria, Victoria, BC, Canada; Dalla Lana School of Public Health, University of Toronto, Toronto, Ontario, Canada

**Keywords:** Sexually Transmitted Diseases, Health Services Utilization, Digital Health, Health Equity, Program Evaluation

## Abstract

**Background:** Digital services for sexually transmitted and blood borne infection (STBBI) testing may influence demand in publicly funded health systems by enabling low barrier, self-directed access to testing, raising concerns about repeated use and sustainability. We examined longitudinal utilization of GetCheckedOnline, British Columbia’s digital STBBI testing service, to characterize testing trajectories and assess factors associated with higher intensity use.

**Methods:** We conducted a retrospective cohort study using GetCheckedOnline program data for users who created an account between April 2020 and November 2022, with 24 months of follow-up. We used group-based trajectory modelling to identify patterns of testing over time among (1) all users and (2) users with at least one test. Multilevel regression models with local health area random intercepts were used to examine associations between higher intensity trajectory membership, individual risk indicators, and geographic clustering.

**Results:** Among 34,228 users, 22,542 (65.9%) completed at least one test and 42,451 tests were conducted (median 1; range 0-44). Two trajectories were identified in both analytic samples, with a minority demonstrating sustained higher intensity testing. The top 10% of users accounted for 39.6% of tests. Higher intensity trajectory membership was associated with sexual risk indicators including having multiple partners, condomless sex with multiple partners, and prior STBBI diagnosis. Geographic clustering across local health areas was modest in the null model (ICC 0.042) and attenuated with adjustment.

**Conclusion:** GetCheckedOnline utilization reflects a prevention-oriented pattern that appears more consistent with service needs than indiscriminate overuse. A small subset of users with elevated sexual risk account for higher-intensity testing. Findings support risk aligned stewardship including education and differentiated guidance, rather than universal restrictions to reducing testing volumes.

**Author Summary:** Digital services are being used to make testing for sexually transmitted and blood borne infections (STBBIs) easier to access. Some health systems decision makers worry that online testing could lead to too much testing and higher costs. We examined how people used GetCheckedOnline, British Columbia’s public online STBBI testing service, over two years. Most people used the service only once or tested occasionally (less than once in six months). A smaller group tested more often. People who tested more often were also more likely to report higher sexual health risk, such as having multiple partners or a recent STBBI diagnosis. This suggests that repeat testing is often done by people who may benefit from testing more often, rather than people testing without need. For health systems, this means it may be more helpful to give clear guidance and education about when to test instead of placing broad limits on access to online testing.

## Introduction

Digital technologies are increasingly used by public health systems to expand access to preventive services while reducing reliance on in-person clinical infrastructure [1, 2]. Among these, digital services for sexually transmitted and blood-borne infection (STBBI) testing have been implemented in several jurisdictions to address barriers related to stigma, geography, costs and limited access to clinical services [2–4]. By enabling self-directed access to laboratory testing, these services are well-suited for asymptomatic screening and have become an important component of current STBBI prevention strategies [5, 6]. Their uptake accelerated during the COVID-19 pandemic alongside broader shifts toward online and virtual models of care [7, 8]. However, the permissive design of some of these publicly-funded testing services raise concerns regarding sustainability and resource stewardship [9]. By circumventing health provider gatekeeping, digital services may increase testing volumes and facilitate repeated testing outside recommended screening intervals [9, 10]. This has prompted debates about induced demand, and potential over-testing, where digital services may disproportionately be used by individuals perceived to be at lower clinical risk, sometimes described as “worried well” testers [9, 11]. Such debates are salient considering currently constrained health system resources and rising laboratory and service delivery costs.

However, empirical evidence regarding the appropriateness and implications of frequent digital STBBI testing remains mixed. Evaluations in the United Kingdom and Europe reveal lower costs per diagnosis and increased systems efficiency as digital testing scales [1, 4]. However, prior work by our research team showed that 28% of GetCheckedOnline testing episodes between 2014 and 2024 occurred more frequently than recommended by Canadian STBBI screening guidelines, but these tests yielded higher positivity than tests conducted at recommended intervals [12]. This suggests that frequent testing may reflect appropriate clinical need, including pre-exposure prophylaxis (PrEP) related monitoring, service design effects, limited access to primary care, or evolving risk profiles, rather than indiscriminate overuse [6, 12]. Understanding these dynamics, particularly in post COVID-19 cohorts, is critical for informing sustainability decisions for the service. Aggregate measures of testing volume alone provide limited insight into how individuals use digital testing services over time. Longitudinal analyses that characterize trajectories of service use and examine how individual, health system, and community contexts influence these trajectories can provide more evidence for policy and service design decisions.

GetCheckedOnline, British Columbia’s publicly funded digital STBBI testing service, provides a unique opportunity to examine these issues at scale [13]. As program partners consider how best to sustain and integrate the service within the provincial health system, evidence is needed to understand who uses the service, under what circumstances, and how these affect resource distribution and system fit [14]. Because digital testing services operate within broader health systems, geographic variation in local service contexts, including differences in primary care attachment and community characteristics, may also influence usage patterns, especially where access to in person care is limited. Therefore, assessing equity in this context requires examining both overall use among registered users and patterns of repeated testing among those who complete tests. Therefore, this study aimed to characterize longitudinal patterns of GetCheckedOnline use and examine how individual demographic and sexual risk factors, as well as health system and community contexts, are associated with distinct testing trajectories. We hypothesized that a small subset of users would account for a disproportionate share of testing volume and that higher intensity testing trajectories would be driven primarily by individual sexual risk and service need (e.g., PrEP monitoring) rather than geographic variation in health system context.

## Results

### Description of the cohort

The study included 34,228 users who created a GetCheckedOnline account between April 1, 2020, and November 30, 2022, with up to 24 months of follow-up. Overall, 22,542 users (65.9%) completed at least one test, while 11,686 (34.1%) did not test during follow-up (Table 1). The mean age was 30 years (SD 9), and users predominantly identified as men (49%) or women (48%) with gender diverse people being the minority (2.9%). Over half of users self-identified as white (53%), although ethnicity data were missing for 20%. Compared with non-testers, users who completed at least one test more frequently identified as men who have sex with men (MSM) exclusively (10.0% vs 6.9%). The proportion of first-time testers was similar among testers and non-testers (19%).

**Table 1:**
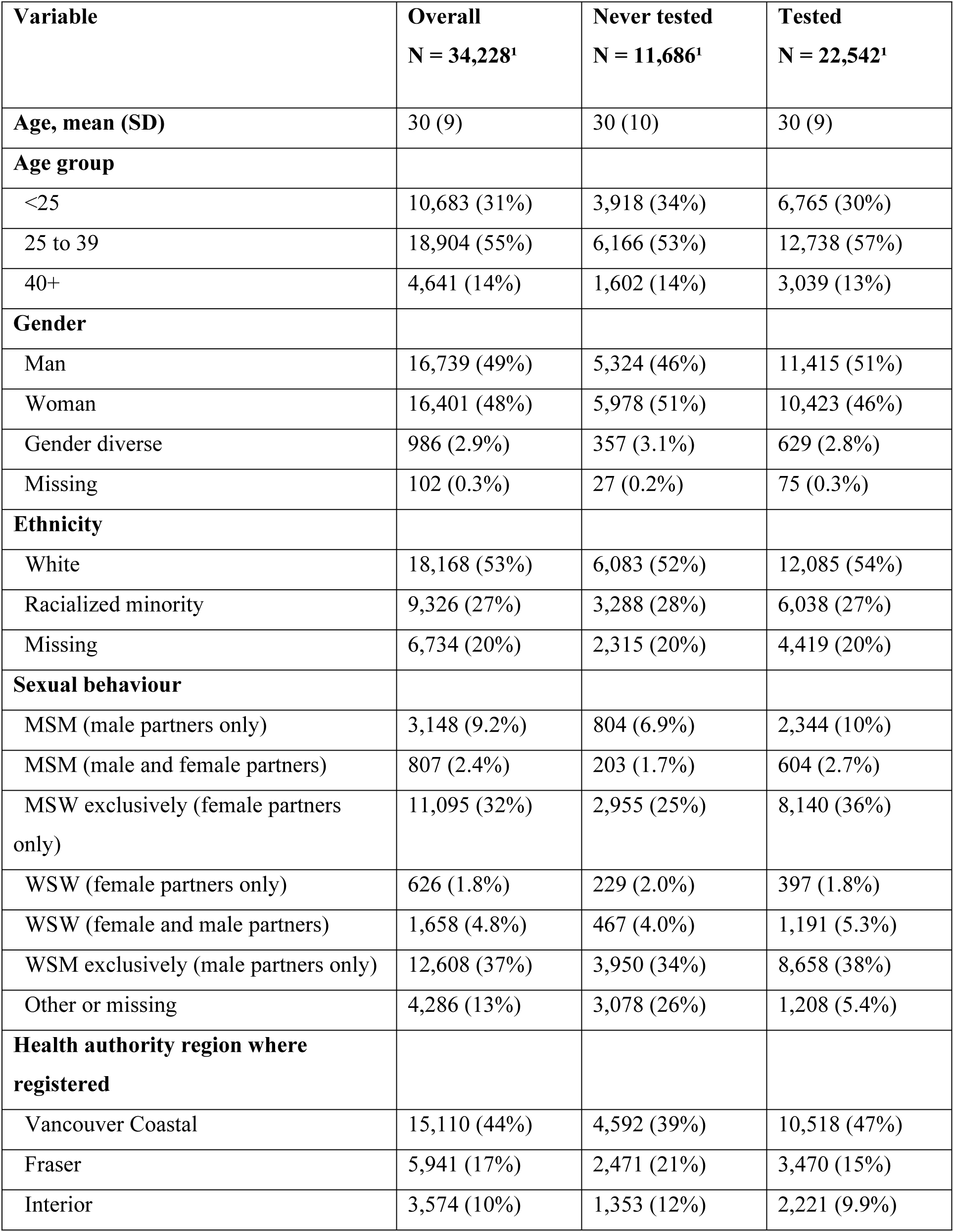
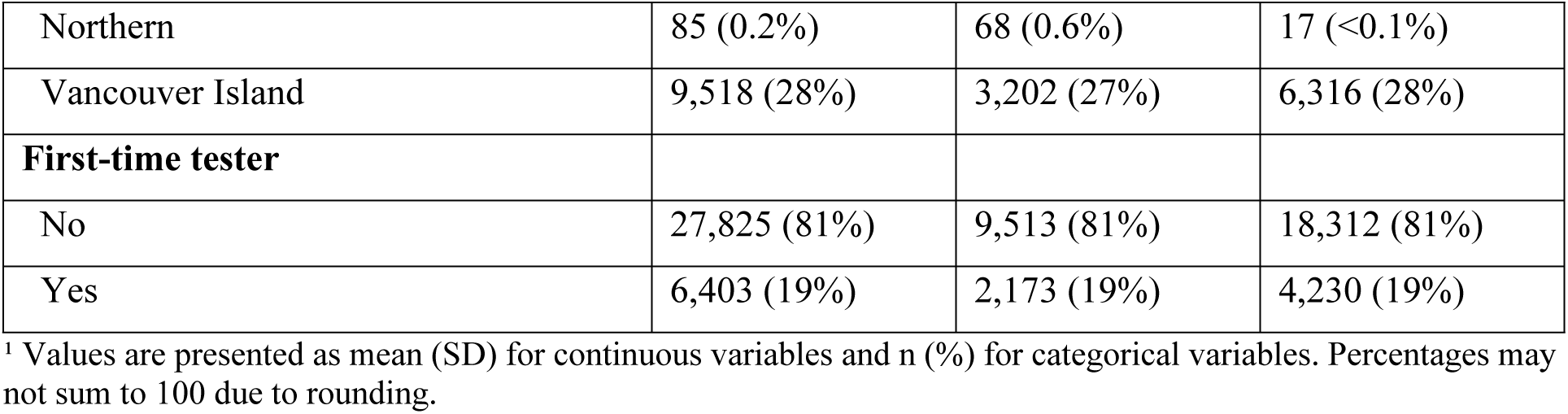
Baseline characteristics of GetCheckedOnline users overall and by testing status (never tested vs tested at least once), April 1, 2020, to November 30, 2022 (N = 34,228)

| **Variable** | **Overall**<br>**N = 34,228<sup>1</sup>** | **Never tested**<br>**N = 11,686<sup>1</sup>** | **Tested**<br>**N = 22,542<sup>1</sup>** |
| --- | --- | --- | --- |
| **Age, mean (SD)** | 30 (9) | 30 (10) | 30 (9) |
| **Age group** |  |  |  |
| <25 | 10,683 (31%) | 3,918 (34%) | 6,765 (30%) |
| 25 to 39 | 18,904 (55%) | 6,166 (53%) | 12,738 (57%) |
| 40+ | 4,641 (14%) | 1,602 (14%) | 3,039 (13%) |
| **Gender** |  |  |  |
| Man | 16,739 (49%) | 5,324 (46%) | 11,415 (51%) |
| Woman | 16,401 (48%) | 5,978 (51%) | 10,423 (46%) |
| Gender diverse | 986 (2.9%) | 357 (3.1%) | 629 (2.8%) |
| Missing | 102 (0.3%) | 27 (0.2%) | 75 (0.3%) |
| **Ethnicity** |  |  |  |
| White | 18,168 (53%) | 6,083 (52%) | 12,085 (54%) |
| Racialized minority | 9,326 (27%) | 3,288 (28%) | 6,038 (27%) |
| Missing | 6,734 (20%) | 2,315 (20%) | 4,419 (20%) |
| **Sexual behaviour** |  |  |  |
| MSM (male partners only) | 3,148 (9.2%) | 804 (6.9%) | 2,344 (10%) |
| MSM (male and female partners) | 807 (2.4%) | 203 (1.7%) | 604 (2.7%) |
| MSW exclusively (female partners only) | 11,095 (32%) | 2,955 (25%) | 8,140 (36%) |
| WSW (female partners only) | 626 (1.8%) | 229 (2.0%) | 397 (1.8%) |
| WSW (female and male partners) | 1,658 (4.8%) | 467 (4.0%) | 1,191 (5.3%) |
| WSM exclusively (male partners only) | 12,608 (37%) | 3,950 (34%) | 8,658 (38%) |
| Other or missing | 4,286 (13%) | 3,078 (26%) | 1,208 (5.4%) |
| **Health authority region where registered** |  |  |  |
| Vancouver Coastal | 15,110 (44%) | 4,592 (39%) | 10,518 (47%) |
| Fraser | 5,941 (17%) | 2,471 (21%) | 3,470 (15%) |
| Interior | 3,574 (10%) | 1,353 (12%) | 2,221 (9.9%) |
| Northern | 85 (0.2%) | 68 (0.6%) | 17 (<0.1%) |
| Vancouver Island | 9,518 (28%) | 3,202 (27%) | 6,316 (28%) |
| **First-time tester** |  |  |  |
| No | 27,825 (81%) | 9,513 (81%) | 18,312 (81%) |
| Yes | 6,403 (19%) | 2,173 (19%) | 4,230 (19%) |

<sup>1</sup> Values are presented as mean (SD) for continuous variables and n (%) for categorical variables. Percentages may not sum to 100 due to rounding.

### Distribution of testing volume

A total of 42,451 tests were completed during follow-up (median per user: 1; range: 0–44; Supplementary Table S1). Over one-third of users (34.1%) did not complete a test. Among all users, testing frequency was highly right-skewed: 90% completed three or fewer tests, 95% completed four or fewer tests, and 99% completed eight or fewer tests. As shown in Figure 1, testing volume was unevenly distributed across users. The bottom 90% of users accounted for 60.4% of completed tests, while the top 10% accounted for 39.6%. Similarly, the top 5% and top 1% of users accounted for 25.6% and 8.3% of tests, respectively.

**Figure 1:**
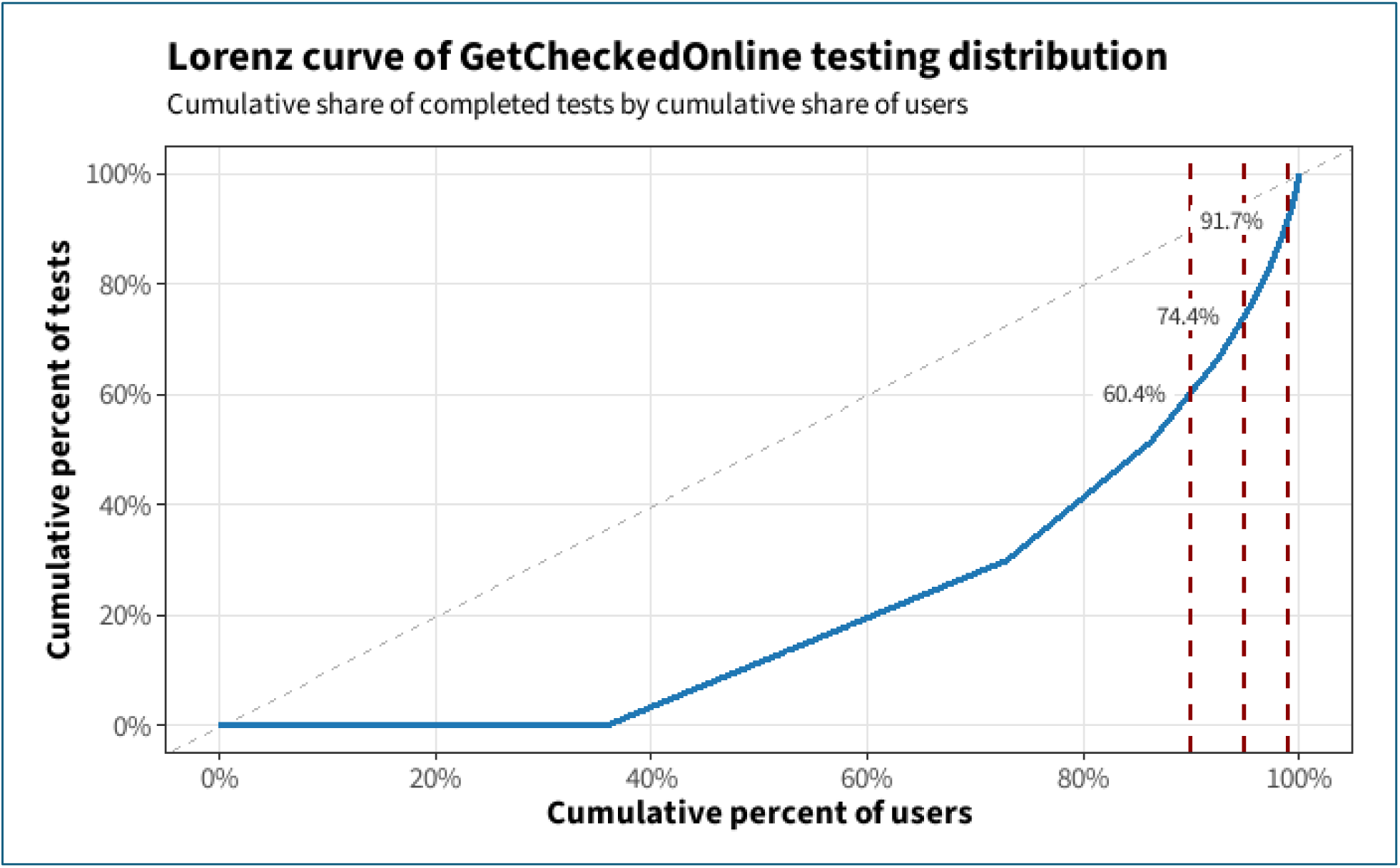
Lorenz curve of completed GetCheckedOnline tests over the 2-year follow-up period. The curve shows the cumulative proportion of completed tests plotted against the cumulative proportion of users, ordered by testing volume. The diagonal line represents perfect equality. Vertical dashed lines indicate the 90th, 95th, and 99th percentiles of users (from left to right).

### Testing trajectories among all users and among users with at least one completed test

Finite mixture Poisson models identified two longitudinal testing trajectories among all users (N = 34,228; Figure 2a). A minority higher-intensity group (n = 4,150; 12.1%) demonstrated sustained repeat testing, averaging approximately 1.4 tests per 6-month interval early after account creation and declining modestly over follow-up. Most users (n = 30,078; 87.9%) exhibited low-intensity testing, with most testing occurring in the first follow-up interval. Restricting analyses to users who completed at least one test (N = 22,542) yielded a similar two-class structure (Figure 2b). A small higher-intensity testing group (n = 2,183; 9.7%) maintained sustained repeat testing across follow-up, while most testers were low intensity, concentrating testing within the first six months. Compared with lower-intensity testers, higher-intensity testers more frequently identified as MSM (20.0% vs 9.4%), had a higher prevalence of prior STBBI diagnoses (41.0% vs 12.0%), and more often reported condomless sex with multiple partners (Table 3).

**Figure 2:**
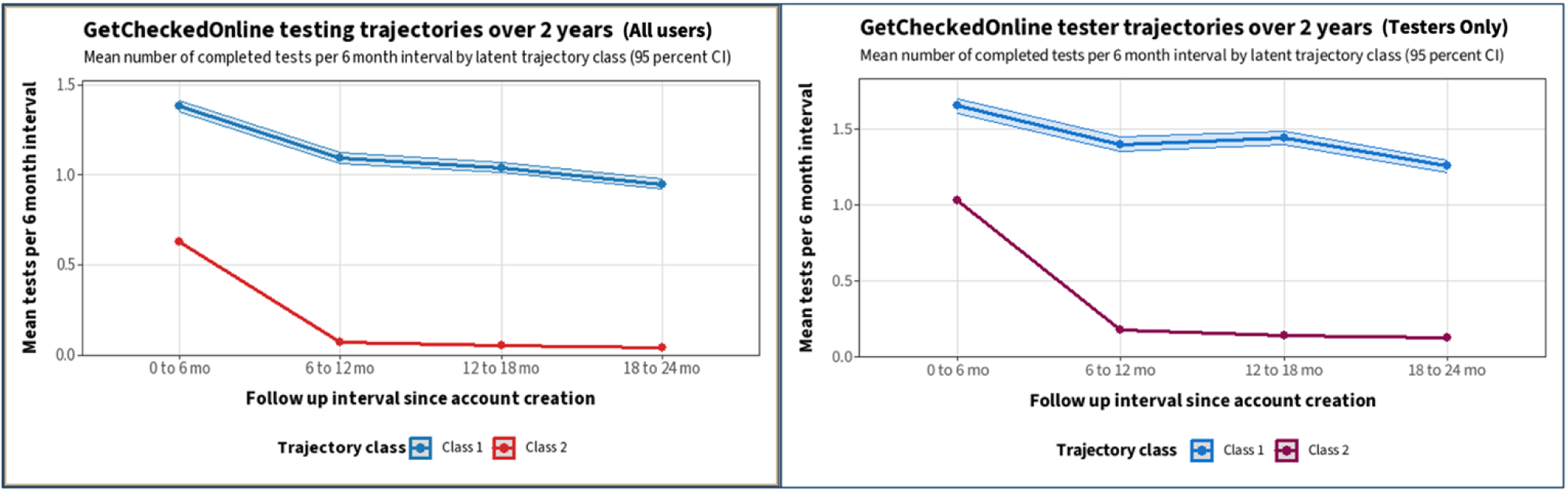
Estimated latent testing trajectories in GetCheckedOnline, a public digital STBBI testing service, over two years of follow up. Panel A shows trajectories among all registered users, and Panel B shows trajectories among users with at least one completed test. Lines represent the estimated mean number of completed tests per user per 6-month interval. Class 1 (blue) represents the sustained higher-intensity testing trajectory, characterized by consistent testing over time. Class 2 (red) represents the lower-intensity or episodic testing trajectory, characterized by an initial test followed by declining utilization. Shaded areas represent 95% confidence intervals.

**Table 2:** Baseline characteristics of GetCheckedOnline users by latent trajectory class.

| **Variable** | **High intensity users (Class 1) (n = 4,150)** | **Lower intensity users (Class 2) (n = 30,078)** | **Overall (n = 34,228)** |
| --- | --- | --- | --- |
| **Age group** |  |  |  |
| <25 years | 1,126 (27%) | 9,557 (32%) | 10,683 (31%) |
| 25 to 39 years | 2,401 (58%) | 16,503 (55%) | 18,904 (55%) |
| 40+ years | 623 (15%) | 4,018 (13%) | 4,641 (14%) |
| **Gender** |  |  |  |
| Man | 2,049 (49%) | 14,690 (49%) | 16,739 (49%) |
| Woman | 1,933 (47%) | 14,468 (48%) | 16,401 (48%) |
| Gender diverse | 150 (3.6%) | 836 (2.8%) | 986 (2.9%) |
| Missing | 18 (0.4%) | 84 (0.3%) | 102 (0.3%) |
| **Ethnicity** |  |  |  |
| White | 2,181 (53%) | 15,987 (53%) | 18,168 (53%) |
| Racialized minority | 1,164 (28%) | 8,162 (27%) | 9,326 (27%) |
| Missing | 805 (19%) | 5,929 (20%) | 6,734 (20%) |
| **Sexual behaviour** |  |  |  |
| MSM (male partners only) | 683 (16%) | 2,465 (8.2%) | 3,148 (9.2%) |
| MSM (male and female partners) | 158 (3.8%) | 649 (2.2%) | 807 (2.4%) |
| MSW exclusively | 1,145 (28%) | 9,950 (33%) | 11,095 (32%) |
| WSW (female partners only) | 68 (1.6%) | 558 (1.9%) | 626 (1.8%) |
| WSW (female and male partners) | 283 (6.8%) | 1,375 (4.6%) | 1,658 (4.8%) |
| WSM exclusively | 1,554 (37%) | 11,054 (37%) | 12,608 (37%) |
| Other or missing | 259 (6.2%) | 4,027 (13%) | 4,286 (13%) |
| **Health authority where registered** |  |  |  |
| Vancouver Coastal | 2,058 (50%) | 13,052 (43%) | 15,110 (44%) |
| Fraser | 672 (16%) | 5,269 (18%) | 5,941 (17%) |
| Interior | 328 (7.9%) | 3,246 (11%) | 3,574 (10%) |
| Northern | 0 (0%) | 85 (0.3%) | 85 (0.2%) |
| Vancouver Island | 1,092 (26%) | 8,426 (28%) | 9,518 (28%) |
| **Primary care attachment** |  |  |  |
| Low (<70%) | 2,912 (70.2%) | 19,212 (63.9%) | 22,124 (64.7%) |
| Medium (70-79%) | 1,055 (25.4%) | 8,899 (29.6%) | 9,954 (29.1%) |
| High (≥80%) | 182 (4.4%) | 1,930 (6.4%) | 2,112 (6.2%) |
| Unknown | 1 (<0.1%) | 37 (0.1%) | 38 (0.1%) |
| **Ethnocultural composition (BCIMD)** |  |  |  |
| Quintile 1 (least deprived) | 100 (2.4%) | 728 (2.4%) | 828 (2.4%) |
| Quintile 2 | 289 (7.0%) | 3,177 (11%) | 3,466 (10%) |
| Quintile 3 | 1,038 (25%) | 7,987 (27%) | 9,025 (26%) |
| Quintile 4 | 2,126 (51%) | 13,521 (45%) | 15,647 (46%) |
| Quintile 5 (most deprived) | 597 (14%) | 4,654 (15%) | 5,251 (15%) |
| Unknown | 0 | 11 | 11 |
| **Economic dependency (BCIMD)** |  |  |  |
| Quintile 1 (least deprived) | 1,911 (46%) | 11,622 (39%) | 13,533 (40%) |
| Quintile 2 | 554 (13%) | 4,540 (15%) | 5,094 (15%) |
| Quintile 3 | 1,487 (36%) | 11,924 (40%) | 13,411 (39%) |
| Quintile 4 | 177 (4.3%) | 1,665 (5.5%) | 1,842 (5.4%) |
| Quintile 5 (most deprived) | 21 (0.5%) | 316 (1.1%) | 337 (1.0%) |
| Unknown | 0 | 11 | 11 |

Percentages may not sum to 100 due to rounding; BCIMD indicates the British Columbia Index of Multiple Deprivation. MSM: Men who have sex with men; MSW: Men who have sex with women; WSM: Women who have sex with men; WSW: Women who have sex with women; Racialized minority: individuals who indicated racial identity other than white – including black, indigenous, people of color etc.

**Table 3:**
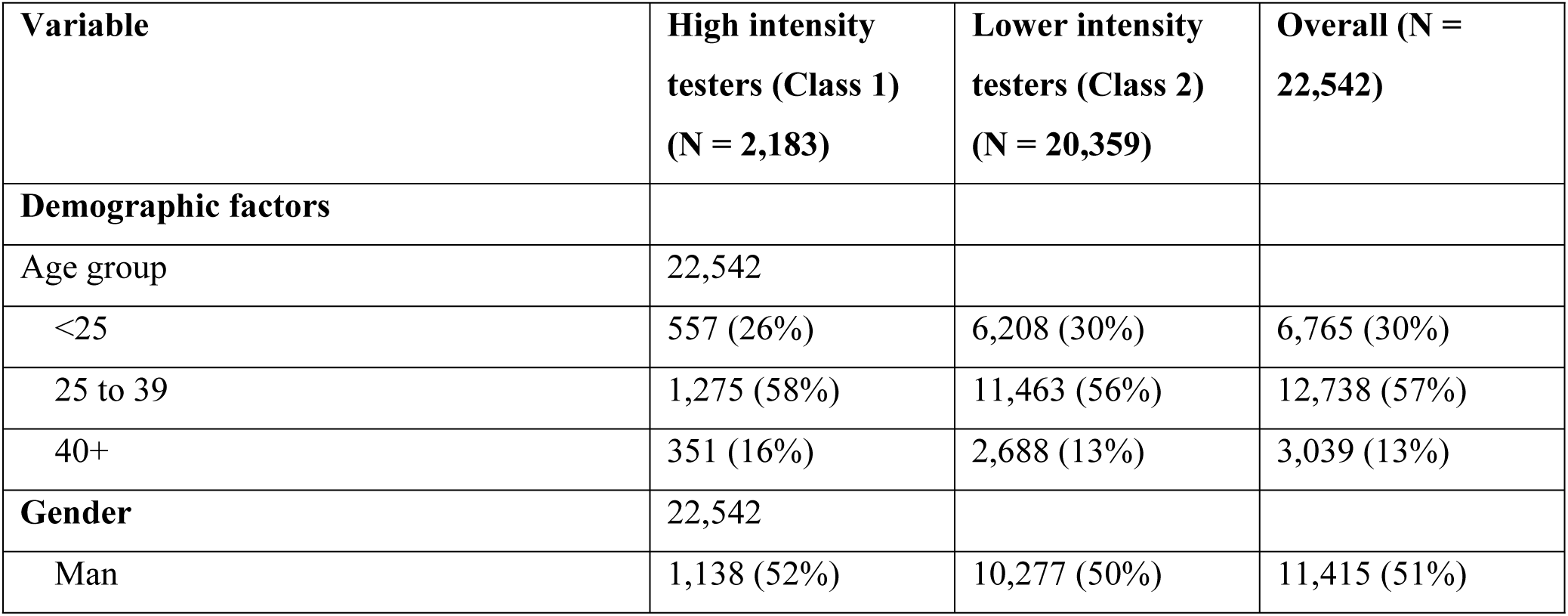
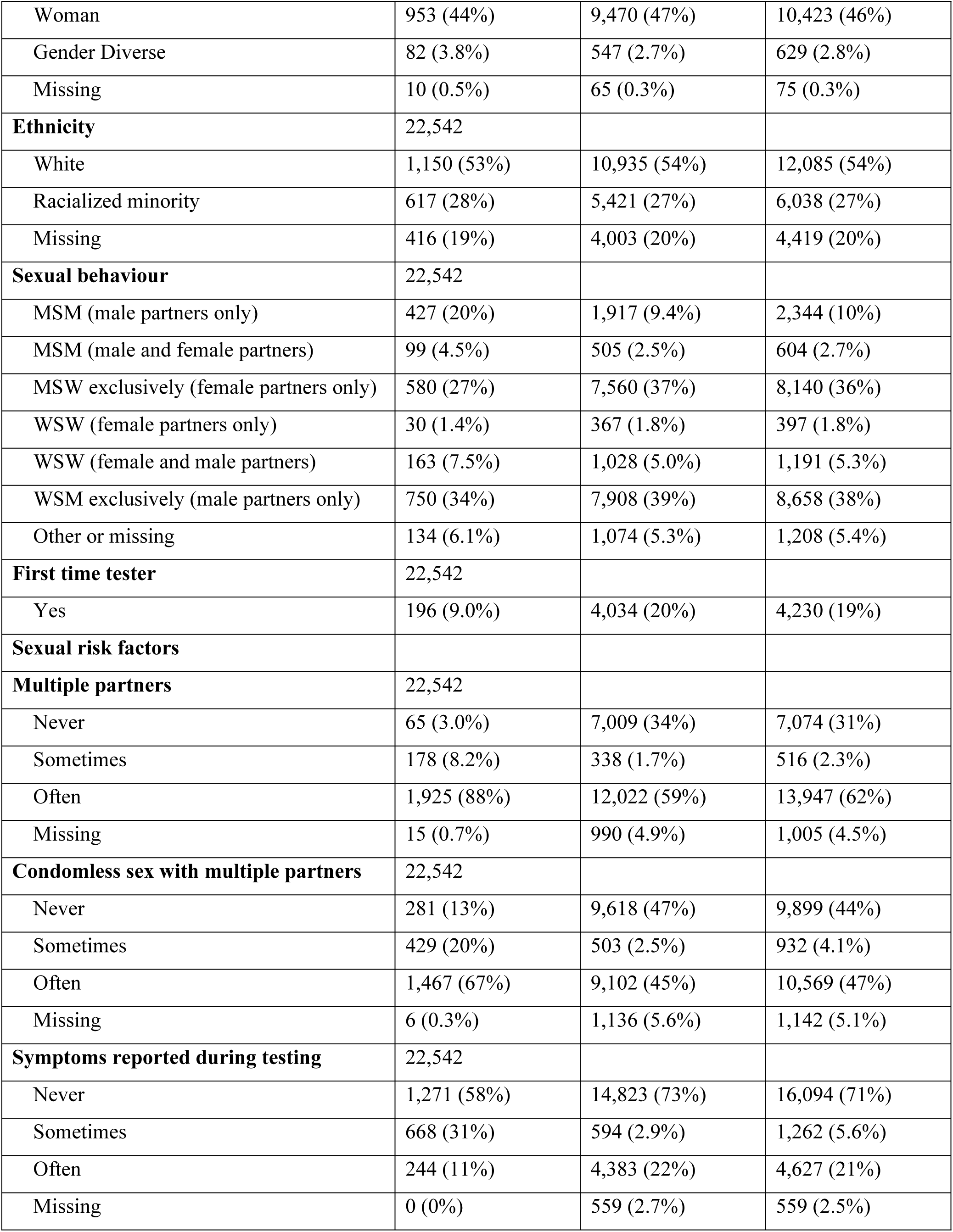
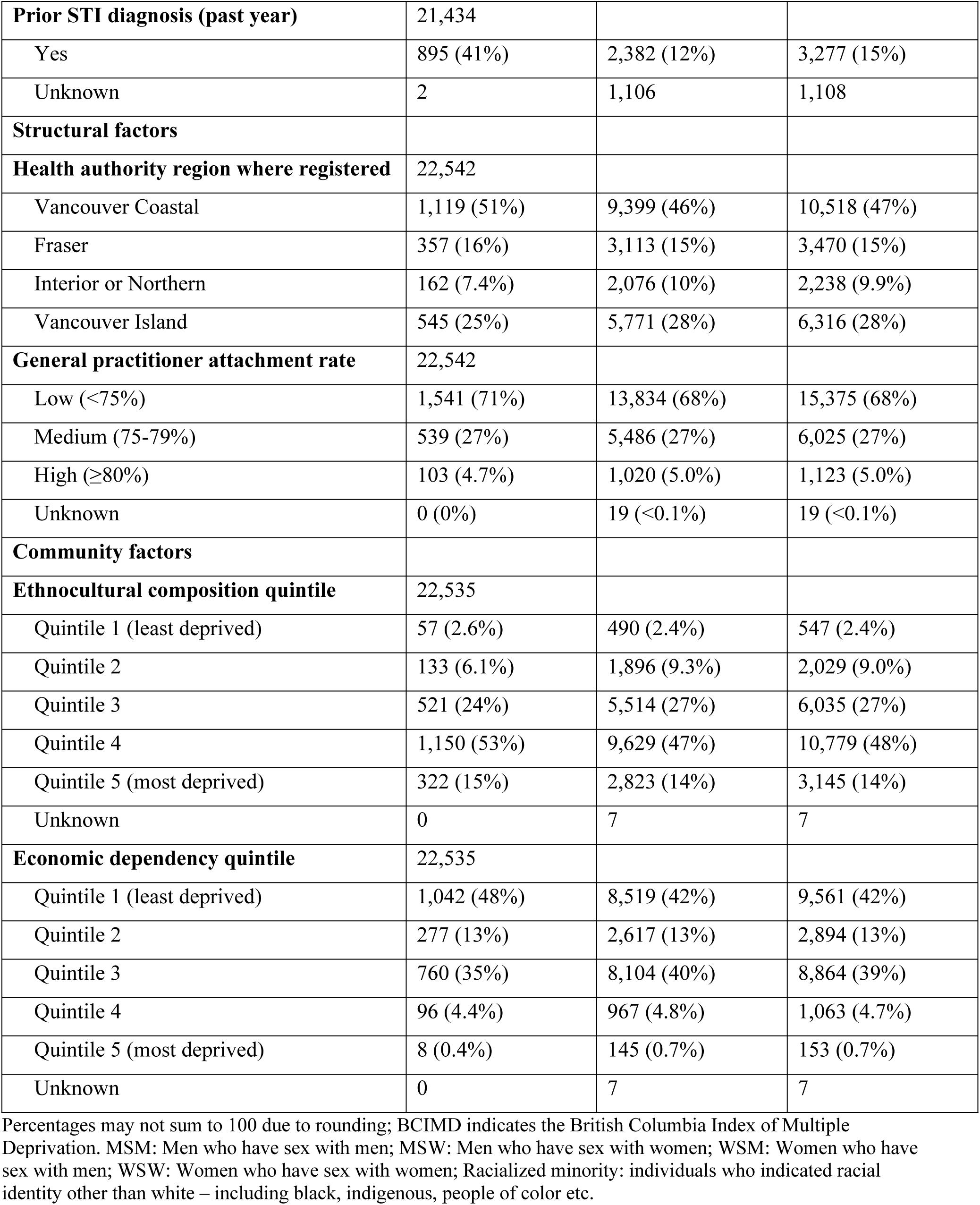
Baseline characteristics of GetCheckedOnline testers by class (N = 22,542)

### Predictors of higher-intensity class membership among all users

In fully adjusted models among all users (Table 4), older age was associated with higher odds of membership in the higher-intensity trajectory compared with age under 25 years (25–39 years: OR 1.19, 95% CI 1.10 to 1.29; ≥40 years: OR 1.38, 95% CI 1.24 to 1.54). Compared with men, women had lower odds of higher-intensity membership (OR 0.51, 95% CI 0.32 to 0.79), while gender-diverse users had higher odds (OR 4.69, 95% CI 3.47 to 6.41). Relative to MSM with male partners only, men who have sex with women exclusively had lower odds of higher-intensity membership (OR 0.43, 95% CI 0.39 to 0.48). Ethnicity, primary care attachment, and most geographic and community-level variables were not significantly associated with trajectory membership, and their inclusion did not meaningfully change model estimates. Geographic clustering of higher-intensity membership was modest (null ICC 0.042) and declined after adjustment for individual and system-level factors (ICC 0.019; Supplementary table S3). Because between-area variance was negligible in fully adjusted models, final models were estimated as fixed-effects logistic regression.

**Table 4:** Adjusted odds ratios for membership in higher intensity trajectory (class 1) among all users.

| Variable | Demographic | Demographic +<br>health system | Demographic +<br>health system +<br>community |
| --- | --- | --- | --- |
| **Age group (ref <25 years)** |  |  |  |
| 25–39 years | 1.20 (1.11, 1.29)*** | 1.20 (1.11, 1.29)*** | 1.19 (1.10, 1.29)*** |
| ≥40 years | 1.38 (1.24, 1.54)*** | 1.38 (1.24, 1.54)*** | 1.38 (1.24, 1.54)*** |
| **Gender (ref Man)** |  |  |  |
| Woman | 0.51 (0.33, 0.78)** | 0.51 (0.33, 0.79)** | 0.51 (0.32, 0.79)** |
| Gender diverse | 4.70 (3.47, 6.37)*** | 4.69 (3.47, 6.35)*** | 4.69 (3.47, 6.41)*** |
| Missing | 5.58 (3.18, 9.81)*** | 5.57 (3.18, 9.76)*** | 5.56 (3.07, 9.66)*** |
| **Ethnicity (ref White)** |  |  |  |
| Racialized minority | 1.02 (0.94, 1.11) | 1.02 (0.94, 1.11) | 1.02 (0.94, 1.11) |
| Missing | 1.02 (0.93, 1.11) | 1.02 (0.93, 1.11) | 1.02 (0.93, 1.11) |
| **Sexual behaviour (ref MSM, male**<br>**partners only)** |  |  |  |
| MSM (male and female partners) | 0.91 (0.75, 1.10) | 0.91 (0.75, 1.10) | 0.90 (0.74, 1.10) |
| MSW exclusively (female partners<br>only) | 0.43 (0.38, 0.48)*** | 0.43 (0.39, 0.48)*** | 0.43 (0.39, 0.48)*** |
| WSW (female partners only) | 0.90 (0.54, 1.50) | 0.90 (0.54, 1.51) | 0.90 (0.54, 1.54) |
| WSW (female and male partners) | 1.56 (0.98, 2.48)† | 1.56 (0.99, 2.48)† | 1.56 (0.98, 2.55)† |
| WSM exclusively (male partners<br>only) | 1.07 (0.68, 1.67) | 1.07 (0.68, 1.67) | 1.07 (0.68, 1.72) |
| Other or missing | 0.14 (0.11, 0.19)*** | 0.14 (0.11, 0.19)*** | 0.14 (0.11, 0.19)*** |
| **Primary care attachment (ref Low**<br>**<75%)** |  |  |  |
| Medium (75-79%) | — | 1.03 (0.67, 1.58) | 1.04 (0.76, 1.43) |
| High (≥80%) | — | 0.90 (0.58, 1.40) | 0.80 (0.52, 1.23) |
| Unknown | — | 0.49 (0.06, 3.97) | 0.58 (0.03, 3.13) |
| **Health authority (ref Vancouver**<br>**Coastal)** |  |  |  |
| Fraser | — | 1.12 (0.73, 1.72) | 1.05 (0.82, 1.34) |
| Interior or Northern | – | 0.81 (0.52, 1.27) | 2.19 (1.01, 5.26)† |
| Vancouver Island | – | 1.03 (0.68, 1.57) | 2.96 (1.42, 6.98)** |
| **Ethnocultural composition**<br>**(BCIMD, ref Quintile 1)** |  |  |  |
| Quintile 2 | – | – | 0.67 (0.52, 0.87)** |
| Quintile 3 | – | – | 0.71 (0.50, 1.01)† |
| Quintile 4 | – | – | 1.82 (0.86, 4.33) |
| Quintile 5 (most deprived) | – | – | 1.96 (0.90, 4.75) |
| **Economic dependency (BCIMD, ref**<br>**Quintile 1)** |  |  |  |
| Quintile 2 | – | – | 0.68 (0.49, 0.93)* |
| Quintile 3 | – | – | 0.74 (0.53, 1.03)† |
| Quintile 4 | – | – | 0.72 (0.42, 1.22) |
| Quintile 5 (most deprived) | – | – | 0.47 (0.23, 0.92)* |

\*Odds ratios (ORs) and 95% confidence intervals (CIs) from stepwise logistic regression models. Class 1 represents the higher-intensity testing trajectory. Reference categories are shown in parentheses. Significance: \*\*\*p<0.001, \*\*p<0.01, p<0.05, †p<0.10. Models were estimated sequentially: demographics only; demographics plus structural factors; and demographics, structural, and community-level factors. Community-level models used fixed-effects logistic regression due to negligible between-area variance. Odds ratios >1 indicate higher odds of membership in Class 1 (higher-intensity testing trajectory).

### Predictors of higher-intensity class membership among users with at least one completed test

Among users with at least one completed test (Table 5), older age remained associated with higher odds of higher-intensity membership (25–39 years: OR 1.31, 95% CI 1.16 to 1.48; ≥40 years: OR 1.74, 95% CI 1.47 to 2.06). Men who have sex with women exclusively had lower odds compared with MSM with male partners only (OR 0.67, 95% CI 0.57 to 0.79). Sexual risk indicators were the strongest predictors of higher-intensity membership. Reporting multiple partners was associated with substantially higher odds (sometimes: OR 9.35, 95% CI 6.66 to 13.23; often: OR 7.19, 95% CI 5.55 to 9.49), as was condomless sex with multiple partners. A prior STBBI diagnosis was also strongly associated with higher-intensity membership (OR 3.10, 95% CI 2.76 to 3.47). Structural and community-level covariates showed smaller and less consistent associations.

**Table 5:**
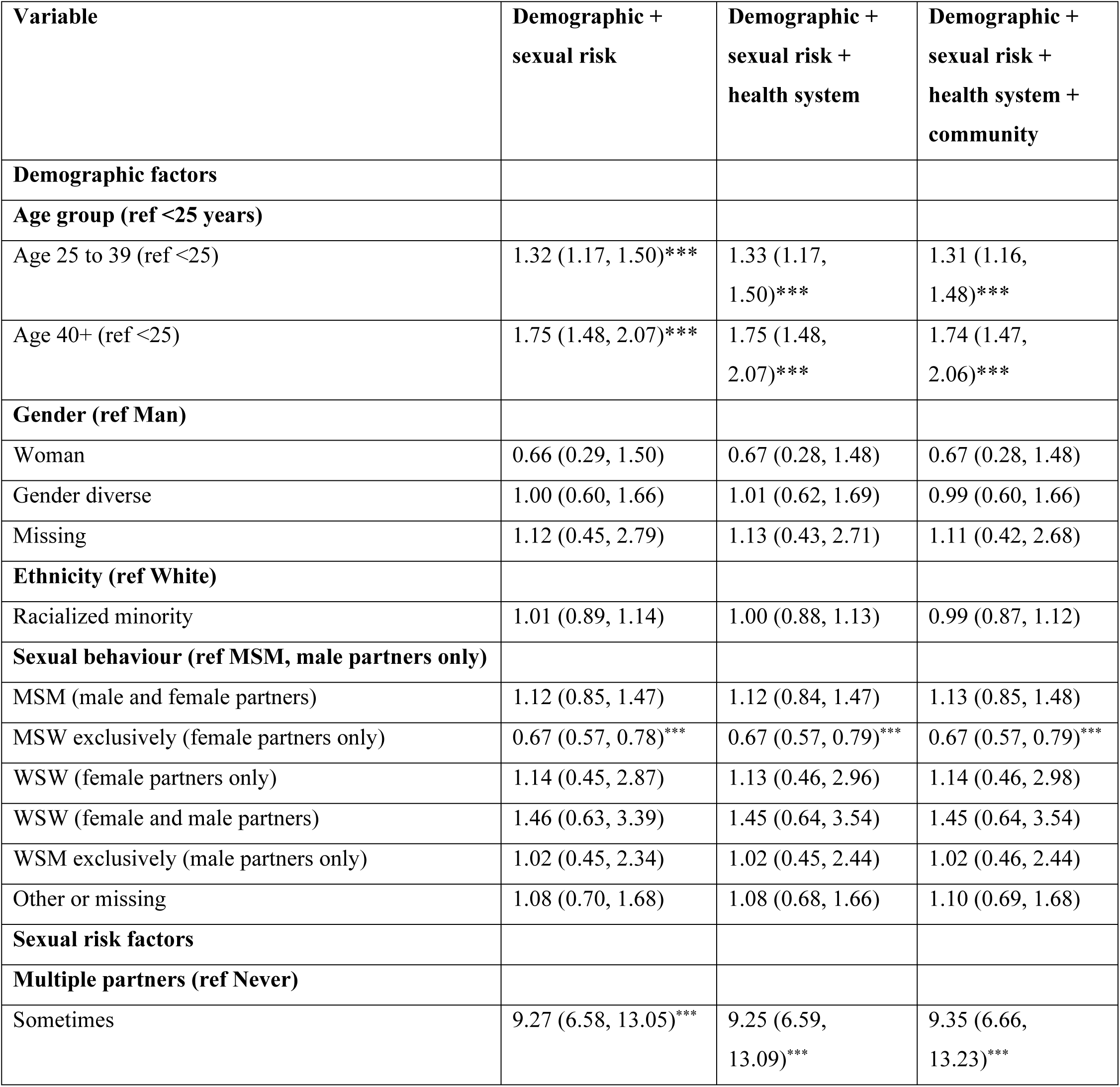
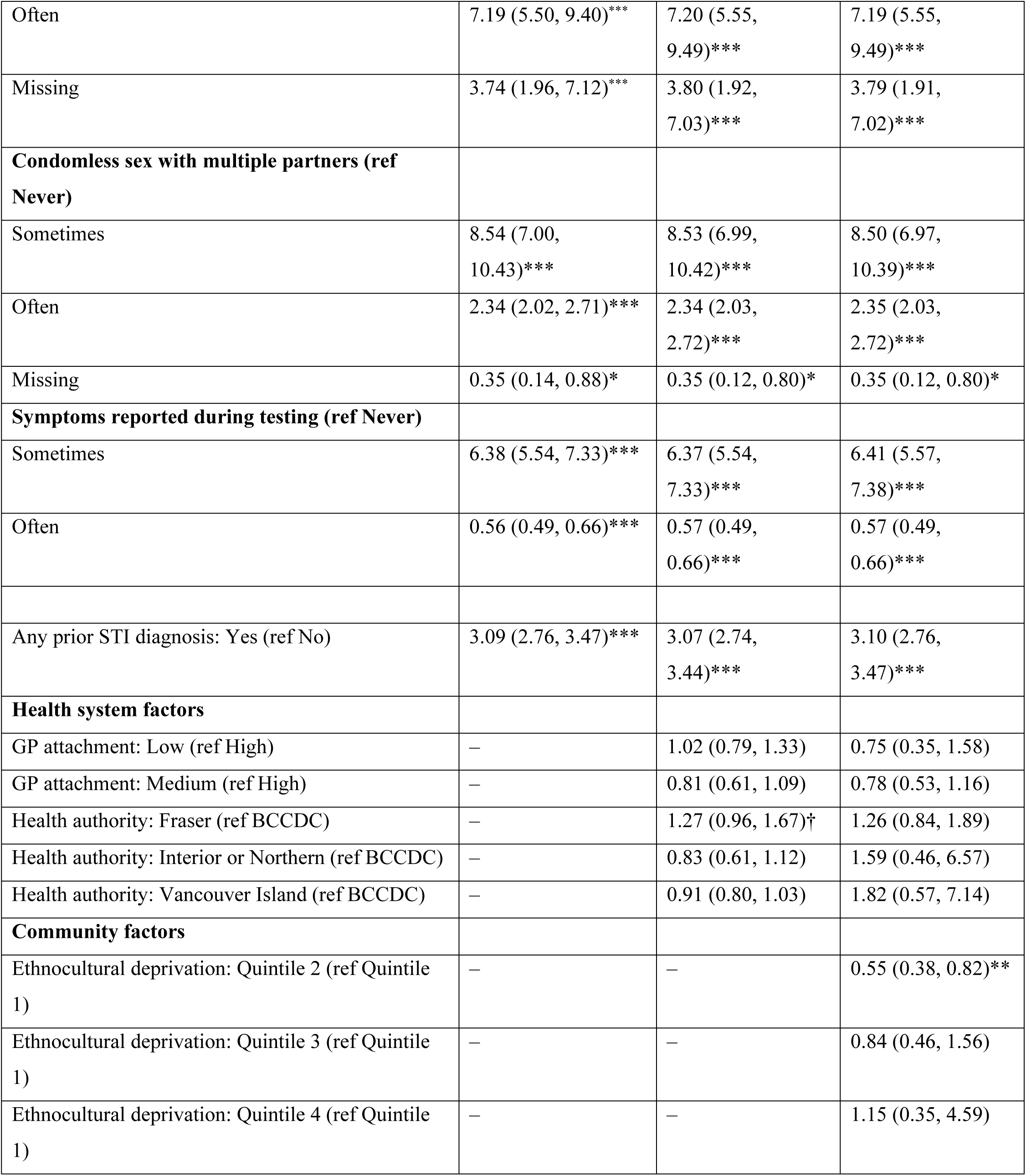
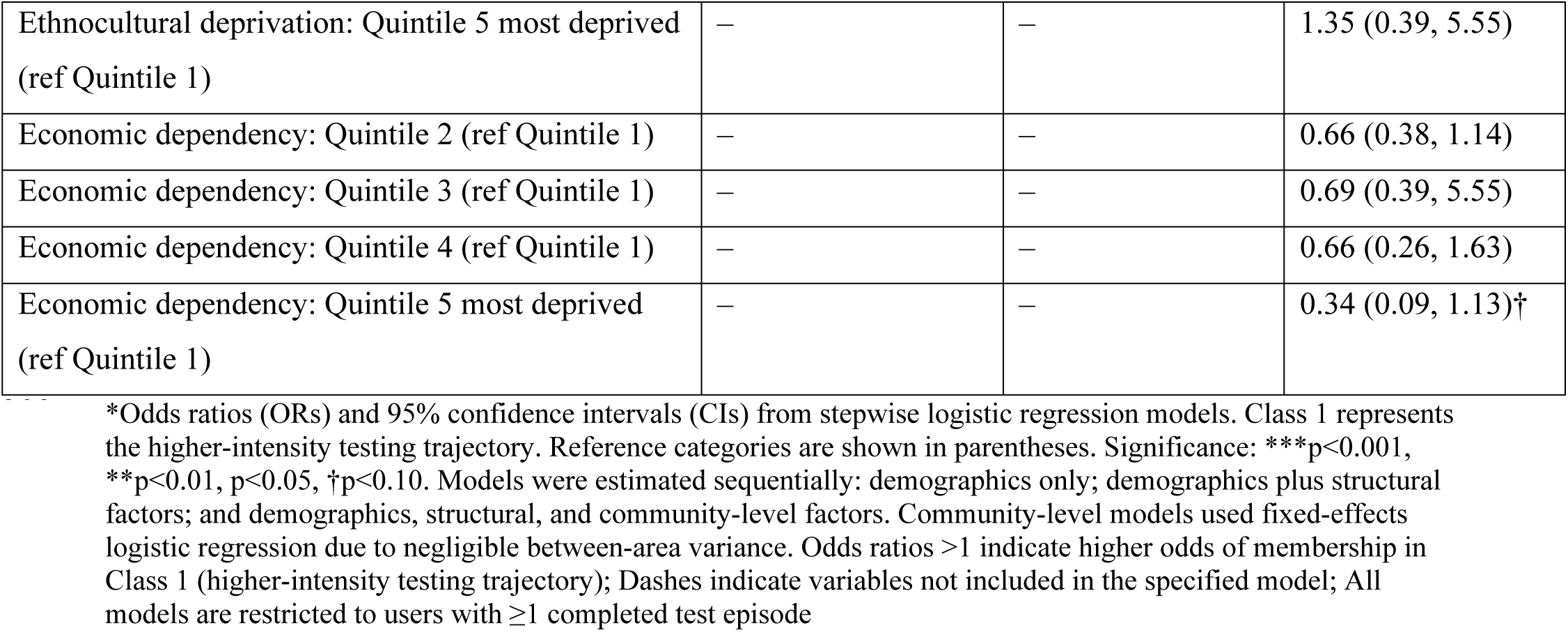
Predictors of class membership among testers: mixed effects and fixed effects logistic regression models showing odds ratios (95% confidence intervals)

### Sensitivity analyses

Sensitivity analyses yielded similar trajectory structures under alternative model specifications. Using 3-month follow-up intervals, three latent testing trajectories were identified among testers (Figure S1 – supplementary material). One small group (4.1%) showed sustained repeat testing across follow-up, the largest group (53.5%) concentrated testing in the first interval with no subsequent use, and a third group (42.5%) demonstrated low-frequency intermittent testing with gradual decline over time. When users at or above the 99th percentile of completed tests were excluded, four latent trajectories were identified (Figure S2 – supplementary material). These included a sustained repeat testing group with lower absolute testing intensity, an early one-time testing group, and two lower-intensity repeat testing groups with declining patterns over follow-up. Across alternative trajectory specifications using linear, quadratic, and cubic time functions, information criteria supported models with additional latent classes; however, class-specific trajectories consistently aligned with higher-intensity repeat testing and lower-intensity testing patterns (Supplementary Table S2).

## Discussion

### Summary of findings

We examined longitudinal patterns of utilization among all registered GetCheckedOnline users and among users who completed at least one test. Across both analytic samples, trajectory modelling identified two dominant patterns: a large group characterized by low-frequency and episodic testing and a smaller group exhibiting sustained higher-frequency testing over time. Analyses including all registered users captured patterns of engagement with the digital service, including individuals who registered but did not proceed to testing, while analyses restricted to testers characterized testing intensity among users who entered the testing pathway. In both samples, membership in the higher-intensity testing trajectory was associated with individual-level sexual risk characteristics, including multiple sexual partners, condomless sex, and prior STBBI diagnoses. In contrast, health system and community-level contextual variables contributed little additional explanatory power, and clustering of testing behaviour at the LHA level was modest and attenuated after adjustment for individual characteristics.

### Comparison with the literature and unique contributions

Most studies of digital STBBI testing services have focused on service uptake, short term outcomes, or aggregate measures of utilization, often using cross-sectional data or short follow up periods [4, 15–17]. While valuable, these approaches limit insight into how individual utilization evolves over time and do not distinguish between sustained need and episodic use [2]. By examining longitudinal testing trajectories over a two-year period, this study provides a more detailed understanding of heterogeneity in digital STBBI testing. Earlier aggregate analyses from our research program suggested over a quarter of tests occurred more frequently than guideline-recommended intervals, raising concerns about potential overuse [12]. The present analysis provides additional context to those concerns by linking patterns of repeat testing to individual level sexual risk indicators. Higher intensity testing was concentrated among users reporting multiple partners, condomless sex with multiple partners, and recent STBBI diagnoses, patterns consistent with current guideline informed testing practices and prevention needs [3, 12]. However, we acknowledge emergent debates about reducing asymptomatic testing frequency especially for chlamydia and gonorrhea to address concerns of overdiagnoses, health systems costs and antimicrobial resistance [18].

Our findings differ from some longitudinal studies of digital STBBI testing in the United Kingdom, which reported pronounced geographic variation in repeat testing and raised concerns about inequitable digital STBBI testing access [15, 16, 19, 20]. In contrast, we observed only modest clustering of higher-intensity testing at the LHA level that diminished after accounting for individual and behavioral factors. These findings indicate that geographic variation in testing volume may reflect differences in population composition and sexual risk patterns rather than structural differences in access to digital STBBI testing services [20]. Consistent with previous studies, higher-intensity testing was more common among MSM [15, 16, 21]. However, unlike studies emphasizing higher uptake among younger users, we found sustained higher-intensity testing to be more strongly associated with older age groups [7]. This pattern may reflect differences in health seeking behaviours and needs including PrEP monitoring, familiarity with testing services, or cumulative exposure to prevention messaging, although these mechanisms require further investigation [22]. Methodologically, this study demonstrates the value of trajectory-based approaches for evaluating digital health interventions. By examining longitudinal patterns of use rather than aggregate service volume, trajectory modelling allows for clearer differentiation between episodic and sustained service engagement and provides a more informative basis for health system planning [23].

### Implications for health systems and service sustainability

Our findings have direct implications for the sustainability of GetCheckedOnline and similar digital STBBI testing services. Repeated testing is often framed as a threat to sustainability due to concerns about demand management and cost containment, including within stewardship initiatives such as Choosing Wisely Canada that emphasize appropriate use of health services [12, 24, 25]. Our study suggests that such concerns require a more nuanced interpretation. Across both analytic samples, sustained high-intensity testing among users with elevated sexual risks imply that repeat testing largely reflects ongoing prevention and care needs rather than indiscriminate or convenience-driven use [12, 17]. Therefore, sustainability strategies may be better supported through targeted, risk-aligned approaches, such as clearer guidance and literacy campaigns on testing intervals and window periods, differentiated testing pathways or integration of self-collection, rather than broad restrictions on testing frequency or geographic access [15, 26]. Such approaches may help support antimicrobial stewardship and resource management while preserving access for individuals who rely on digital testing as part of routine sexual health care. Blanket limitations risk disproportionately affecting users who rely on digital testing as a routine component of their sexual health care [27, 28]. From a health systems perspective, these findings support a view of sustainability as the capacity of digital STBBI services to adaptively meet population needs over time, rather than as static control of service volume [14]. To inform service adaptations, further economic and population-level modeling and evaluation is needed. While existing evidence on cost per diagnosis is encouraging, additional analyses examining system-level impacts, including potential costs avoided and reduction in population-level prevalence through substitution of digital testing for in-person clinical care among higher-intensity users, would strengthen the economic case for these services [1, 18, 29].

Beyond GetCheckedOnline, this study contributes to broader debates on demand induction and de-implementation in digital STBBI testing, where concerns have been raised that expanded access may increase testing frequency, with potential downstream implications for overtreatment and antimicrobial stewardship [9, 25, 30, 31]. Evaluations based solely on aggregate utilization or cost-per-test risk obscuring meaningful heterogeneity in use. Our findings indicate that repeat testing should not be interpreted as inefficient by default but assessed in relation to user sexual risk profiles and longitudinal utilization patterns. Incorporating behavioural and risk-related measures into evaluations is therefore critical to avoid misattributing variation in use to service design or inequitable access [27]. More broadly, this study demonstrates the value of trajectory-based cohort approaches for evaluating digital health interventions by capturing dynamic patterns of use that reflect changing individual risk over time [23, 32]. By moving beyond short-term uptake or binary indicators of use, such methods provide a clearer basis for understanding episodic versus sustained need and for informing equity-oriented and sustainability-focused service planning.

### Future research

Future research should build on these findings by integrating trajectory-based classifications into prospective evaluations of digital STBBI testing services. Linking longitudinal use patterns to outcomes such as diagnoses, downstream care utilization, and costs would enable more robust assessments of value and sustainability [1, 12, 33]. Comparative analyses across jurisdictions and service models would help determine the generalizability of observed trajectories and inform context-sensitive adaptation. Finally, embedding trajectory-informed analyses within economic and implementation studies could support equity-oriented service planning while avoiding blunt demand-reduction approaches.

### Strengths and limitations

This study draws on a large, population level longitudinal dataset and applies trajectory based analytic methods to characterize patterns of digital STBBI testing over time. The consistency of findings across multiple model specifications strengthens confidence in the robustness of the results. Several limitations should be noted. Sexual risk indicators were derived from self-reported data and summarized at the user level, which may introduce measurement error, although these responses reflect information routinely collected as part of service delivery. Geographic clustering was assessed at the LHA and CHSA level, and factors operating at other contextual scales could not be examined. As an observational study conducted within a single publicly funded program, the findings may not be directly generalizable to privately operated or direct to consumer digital testing models. In addition, the analysis captures testing through GetCheckedOnline only and does not account for concurrent testing through other clinical services. Previous research suggests that only about half of users test exclusively through GetCheckedOnline, while up to 15% test equally or more often through other providers [2]. Therefore, overall testing activity across the health system is likely underestimated in this dataset. Further research is needed to identify which users test concurrently across services and whether they overlap with the higher intensity testers identified in our study.

To conclude, utilization of GetCheckedOnline follows a clear prevention-oriented pattern: most users test infrequently, while a smaller subset with elevated sexual risk accounts for sustained higher-frequency testing. These findings challenge assumptions that repeat testing within digital STBBI services primarily reflects inefficiency or inappropriate demand and instead suggest that higher-frequency utilization represents appropriate, prevention-oriented care for users with ongoing risk. Considering both registered users and active testers highlights that digital STBBI services serve both as low-barrier entry points for episodic testing and as ongoing prevention tools for individuals with sustained risk. For health systems, this underscores the importance of moving away from blunt volume-based restrictions as has been implemented in other jurisdictions and toward targeted stewardship approaches that preserve access for users with ongoing sexual health needs while supporting the long-term sustainability of digital STBBI testing services.

## Materials and Methods

### Study design and setting

This was a retrospective cohort study assessing users of GetCheckedOnline, which was launched in 2014 and is operated by the British Columbia Centre for Disease Control (BCCDC) in partnership with the BC Public Health Laboratory [13]. The service enables asymptomatic users to access STBBI testing and results online without a clinical visit. Described in detail elsewhere, users create an online account, complete a standardized sexual risk assessment, and receive test recommendations [3, 13]. Specimen collection occurs at participating sites operated by a contracted private laboratory provider, and test results are delivered online. Public health nurses provide follow-up for users with positive results, including treatment referral where indicated. GetCheckedOnline offers testing for chlamydia, gonorrhea, syphilis, HIV, and hepatitis C. During the study period (2022), the service operated in nine urban, suburban, and rural communities across British Columbia, conducting more than 30,000 tests annually, accounting for over 7.5% of all STBBI tests in regions where it operates [3, 7].

### Data sources and study population

We used routinely collected administrative and contextual data linked at the individual, health system, and community levels. Individual-level data were obtained from the GetCheckedOnline program database and included demographic characteristics reported at account creation (age, gender, and race or ethnicity). The database includes sexual risk information collected through a questionnaire completed before each testing episode, enabling characterization of risk related testing behaviours over time. Longitudinal testing activity was derived from program records with completed testing episodes identified using a flag indicating successful specimen collection and testing by the laboratory. These records were used to construct user-level testing trajectories following account creation. Health system and community-level contextual data were obtained from external sources. Primary care access was characterized using area-level estimates of attachment to a general practitioner obtained from the British Columbia Ministry of Health [34]. Community socioeconomic context was captured using the 2021 British Columbia Indices of Multiple Deprivation (BCIMD) [35]. Data were linked using jurisdiction codes to non-identifying service and geographic indicators at the Local Health Area (LHA) and Community Health Service Area (CHSA) level, administrative units used by the British Columbia Ministry of Health to organize health services. The analytic dataset contained no direct personal identifiers. The study population included all individuals who created a GetCheckedOnline account between April 1, 2020, and November 30, 2022, allowing up to two years of follow-up. All users were eligible for inclusion in trajectory analyses, regardless of whether they completed a test. Users with incomplete registration data required for contextual linkage were excluded. Follow up time from account creation was truncated at 24 months to ensure a consistent observation window.

### Measures

The primary outcome was longitudinal GetCheckedOnline testing behaviour, defined using dates of account creation and completed testing episodes. Individual-level covariates included age, gender, and race or ethnicity, measured at account creation. Age was categorized as <25 years (reference), 25–39 years, and ≥40 years. Gender was categorized as man, woman, gender diverse, or missing. Sexual behaviour was categorized as men who have sex with men with male partners only (MSM), MSM with male and female partners, men who have sex with women exclusively (MSW), women who have sex with women exclusively (WSW), women who have sex with women and men, women who have sex with men exclusively (WSM), and other or missing. Sexual risk indicators reported before each testing episode were summarized as the proportion of tests reporting the behavior and categorized as never (0%), sometimes (>0 to <50%), or often (≥50%). Health system context was captured using general practitioner (GP) attachment rates at the CHSA level, categorized as low (<75%), medium (75–79%), and high (≥80%). Community-level socioeconomic context was measured using quintiles of the 2021 British Columbia Indices of Multiple Deprivation at the LHA level [35]. Area-level measures were used as proxies for health service access and community context and were not intended to represent individual-level exposures. For all covariates, a missing or unknown category was included where applicable to retain observations with incomplete data and to minimize bias related to complete-case analysis. All measures were defined a priori based on their relevance to service access, utilization patterns, and sustainability considerations [36, 37].

### Data analysis

We used descriptive analyses to characterize GetCheckedOnline users overall and by testing status (never tested vs tested at least once). To assess how testing volume was distributed across users, we plotted a Lorenz curve and created descriptive summaries of the number of tests completed per user. No formal hypothesis testing was conducted for these descriptive analyses. We used group-based trajectory modeling to identify distinct longitudinal patterns of use [23]. Trajectories were estimated using finite mixture models with a Poisson regression specification for the count of completed tests per follow up interval. We fit models in two analytic samples: (1) all registered users, including users with no completed tests, and (2) users who completed at least one test during follow up. The first sample captures the full spectrum service utilization, including account creation without testing, which is relevant for understanding system-level demand and potential access barriers while the second sample allowed clearer characterization of testing intensity once initial uptake has occurred. For each sample, we created a longitudinal person interval dataset after account creation in which the outcome was the number of completed tests in each interval, allowing zero counts. Time since account creation was discretized into fixed 6 month follow up intervals, reflecting the upper bound of Canadian STBBI screening recommendations for higher risk populations while reducing data sparsity [38, 39]. Polynomial terms for time were included to allow non-linear patterns in testing over time. We estimated models with increasing numbers of latent trajectory groups. Model selection was guided by Bayesian Information Criterion (BIC) values, minimum class size, average posterior probabilities, and substantive interpretability of the identified trajectories [23, 40]. Each user was assigned to the trajectory group corresponding to their highest posterior probability. After trajectory classification, we examined associations between individual level, health system, and community level characteristics and trajectory group membership. Trajectory membership was modeled using multilevel hierarchical logistic regression with random intercepts for LHAs to account for geographic clustering, with models estimated separately for the all-user sample and the tester only sample. When between area variance was negligible or models resulted in singular fits, models were refit as fixed-effects logistic regressions. Contextual variables capturing health system and community characteristics were added sequentially to assess whether patterns of trajectory membership varied across service contexts. Robust standard errors clustered by LHA were used where appropriate. Results are presented as odds ratios with 95 percent confidence intervals (95% CI). All analyses were exploratory and intended to characterize heterogeneity in utilization patterns rather than estimate causal effects. Missing data were handled using missing or unknown categories; no imputation was performed due to the computationally intense nature of finite mixture modelling on such a large dataset [41]. All analyses were conducted using R version 4.4.2 [42].

### Sensitivity analyses

To assess the robustness of the identified testing trajectories, we conducted three sensitivity analyses. First, we re-estimated trajectory models using alternative time discretization, including 3-month follow up intervals, to evaluate sensitivity to the choice of time scale. Second, we examined alternative trajectory specifications by varying the functional form of time (linear, quadratic, and cubic) and the number of latent classes to assess the stability and interpretability of trajectory solutions. Third, to evaluate the influence of extreme testing behavior, we re-estimated trajectory models after excluding users whose total number of completed tests fell at or above the 99th percentile.

### Ethical considerations

This study was approved by the University of British Columbia Behavioural Research Ethics Board (H24-01575) and conducted in accordance with the Declaration of Helsinki. It involved secondary analysis of routinely collected health data and did not require informed consent. Data access was approved by the British Columbia Centre for Disease Control Data Governance Committee. Reporting follows the STROBE guidelines for observational studies [43].

## Acknowledgements

The authors would like to thank Heather Pedersen and Ashley Bentley for their guidance and thoughtful input throughout the project. We also acknowledge colleagues within the BCCDC Clinical Prevention Services team for their support and collaboration.

Finally, we acknowledge the users of GetCheckedOnline, whose engagement with the service and efforts to access testing shape the broader context in which this research was conducted.

## Ethical approval

This study was approved by the University of British Columbia Behavioural Research Ethics Board (H24-01575) and conducted in accordance with the Declaration of Helsinki. It involved secondary analysis of routinely collected health data and did not require informed consent. Data access was approved by the British Columbia Centre for Disease Control Data Governance Committee.

## Declaration of Generative AI in Scientific Writing

During the preparation of this manuscript, the authors used generative artificial intelligence (OpenAI GPT-5.2) to assist with improving the clarity and readability of the text. All outputs were reviewed and edited by the authors, who take full responsibility for the final content of the manuscript.

## Data Availability Statement

The data used in this study are not publicly available due to privacy and data governance restrictions imposed by the British Columbia Centre for Disease Control (BCCDC). The data contain potentially identifiable health information and are subject to institutional and provincial privacy regulations. Access to de-identified data may be considered on reasonable request, subject to approval by the BCCDC Data Governance Committee and relevant ethics approvals.

## Author Contributions

Ihoghosa Iyamu: Conceptualization, Methodology, Formal analysis, Data curation, Writing – original draft, Writing – review & editing.

Devon Haag: Methodology, Supervision, Writing – review & editing.

Sofia Bartlett: Methodology, Supervision, Writing – review & editing.

Catherine Worthington: Supervision, Writing – review & editing.

Daniel Grace: Supervision, Writing – review & editing.

Mark Gilbert: Conceptualization, Data curation, Supervision, Funding acquisition, Writing – review & editing.

## Appendix

### Appendix 1 STROBE Statement—Checklist of items that should be included in reports of cohort studies

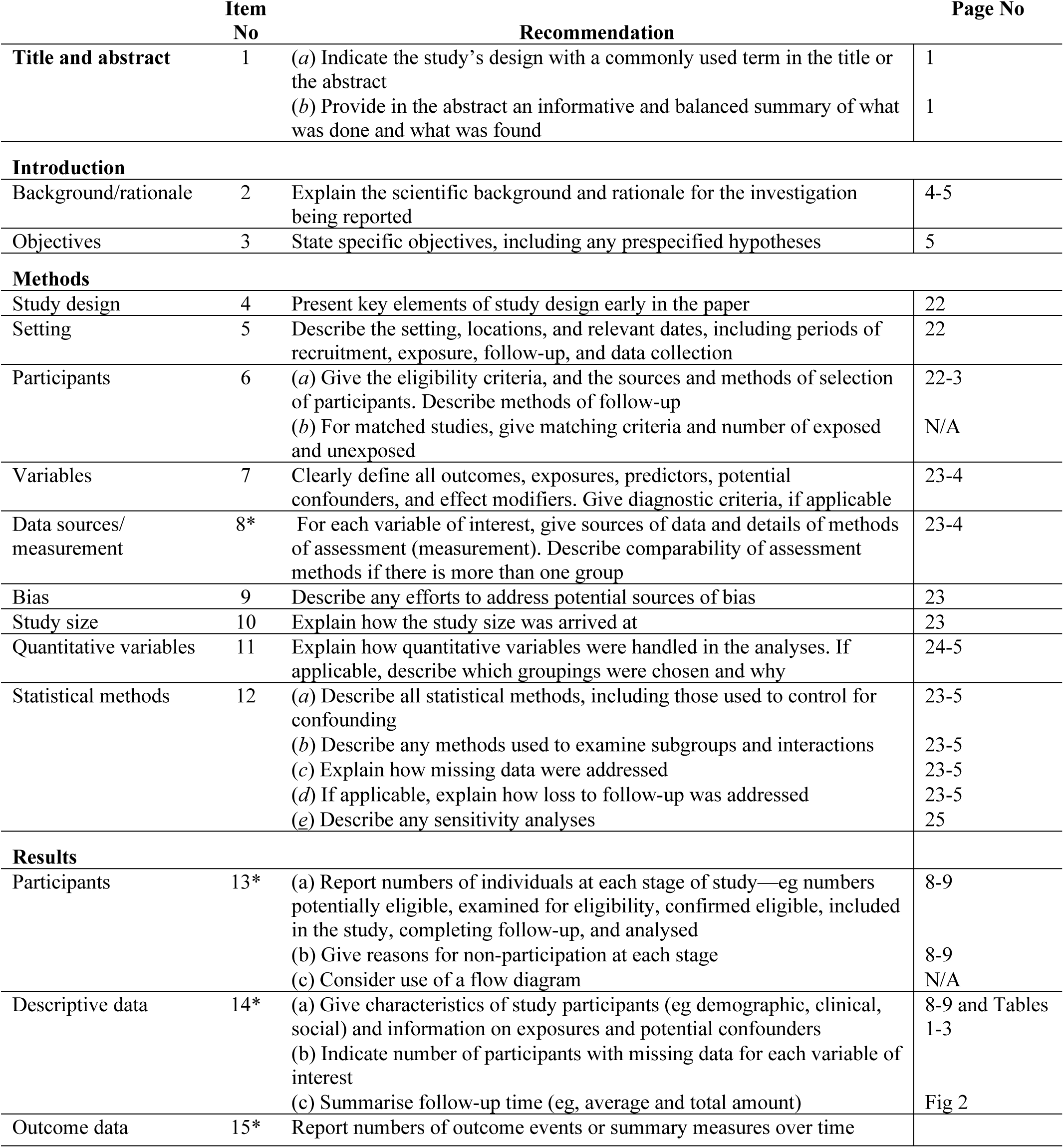
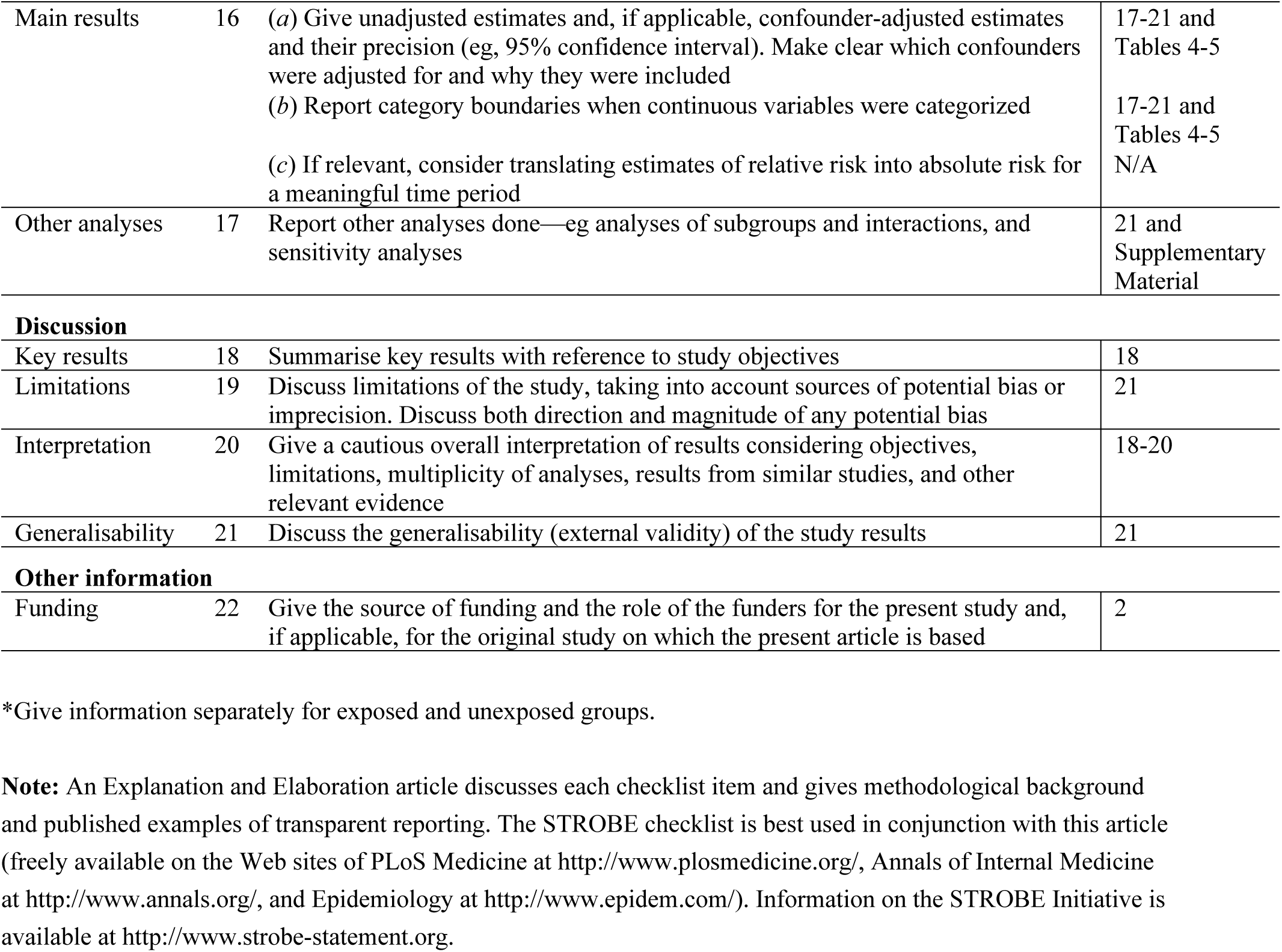

**Supplementary Table S1:**
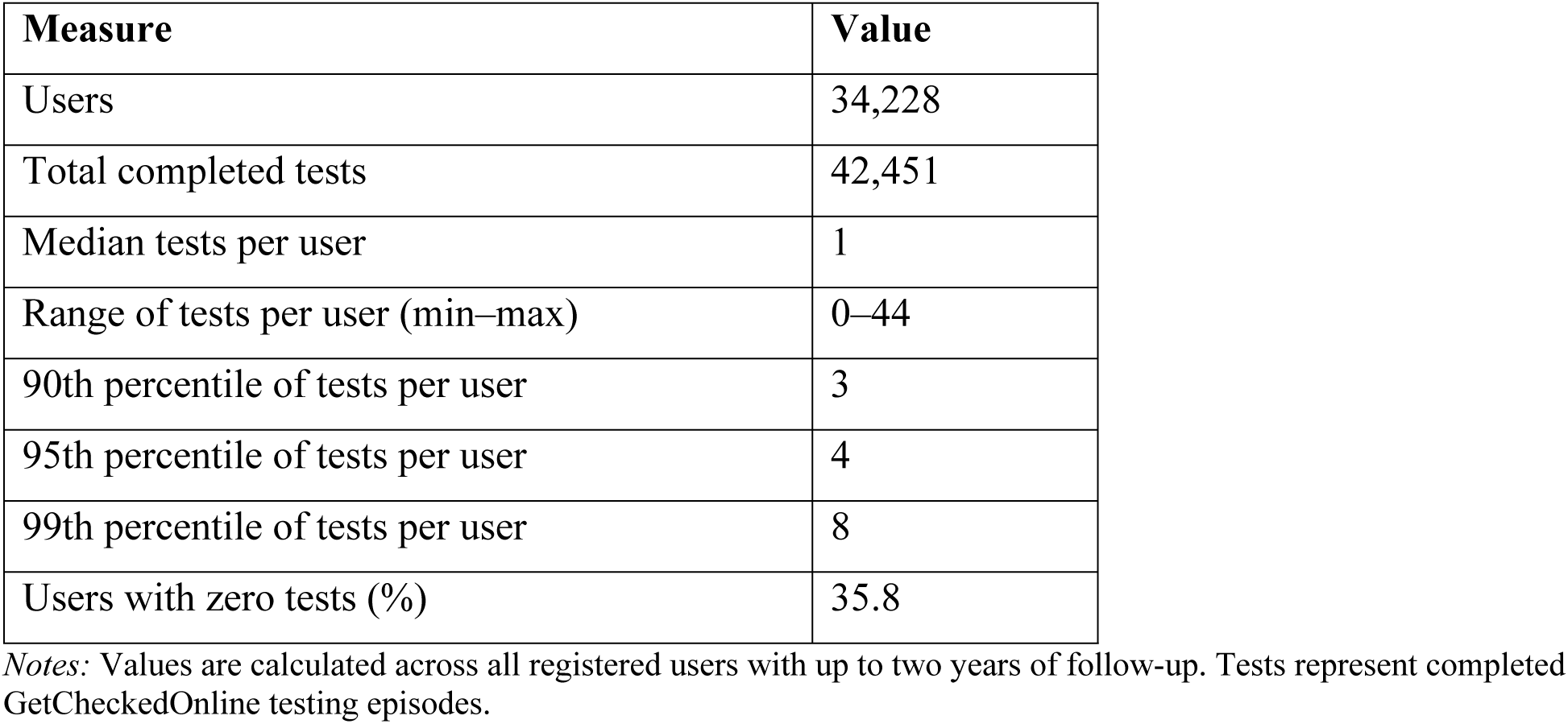
Distribution of completed GetCheckedOnline tests over the 2-year follow-up period.

To further characterize the distribution of testing activity underlying the trajectory analyses, we summarized the total number of completed GetCheckedOnline tests across all registered users with up to two years of follow-up (Table S1). Among 34,228 users, a total of 42,451 tests were completed during the observation period. The median number of tests per user was one, with a wide but highly skewed distribution ranging from zero to 44 tests. Over one third of registered users (35.8%) did not complete any tests during follow-up.

Testing activity was concentrated among a relatively small subset of users. The 90th percentile of users completed three or fewer tests, the 95th percentile completed four or fewer tests, and the 99th percentile completed eight or fewer tests over two years. These summary statistics corroborate the Lorenz curve analysis, demonstrating substantial inequality in testing volume and reinforcing the presence of a small group of higher-frequency testers alongside a much larger group of low-frequency or non-testing users.

### Sensitivity analyses

#### Sensitivity analyses 1 – Group-based trajectory modeling with 3-month discrete time intervals

**Supplementary Figure S1:**
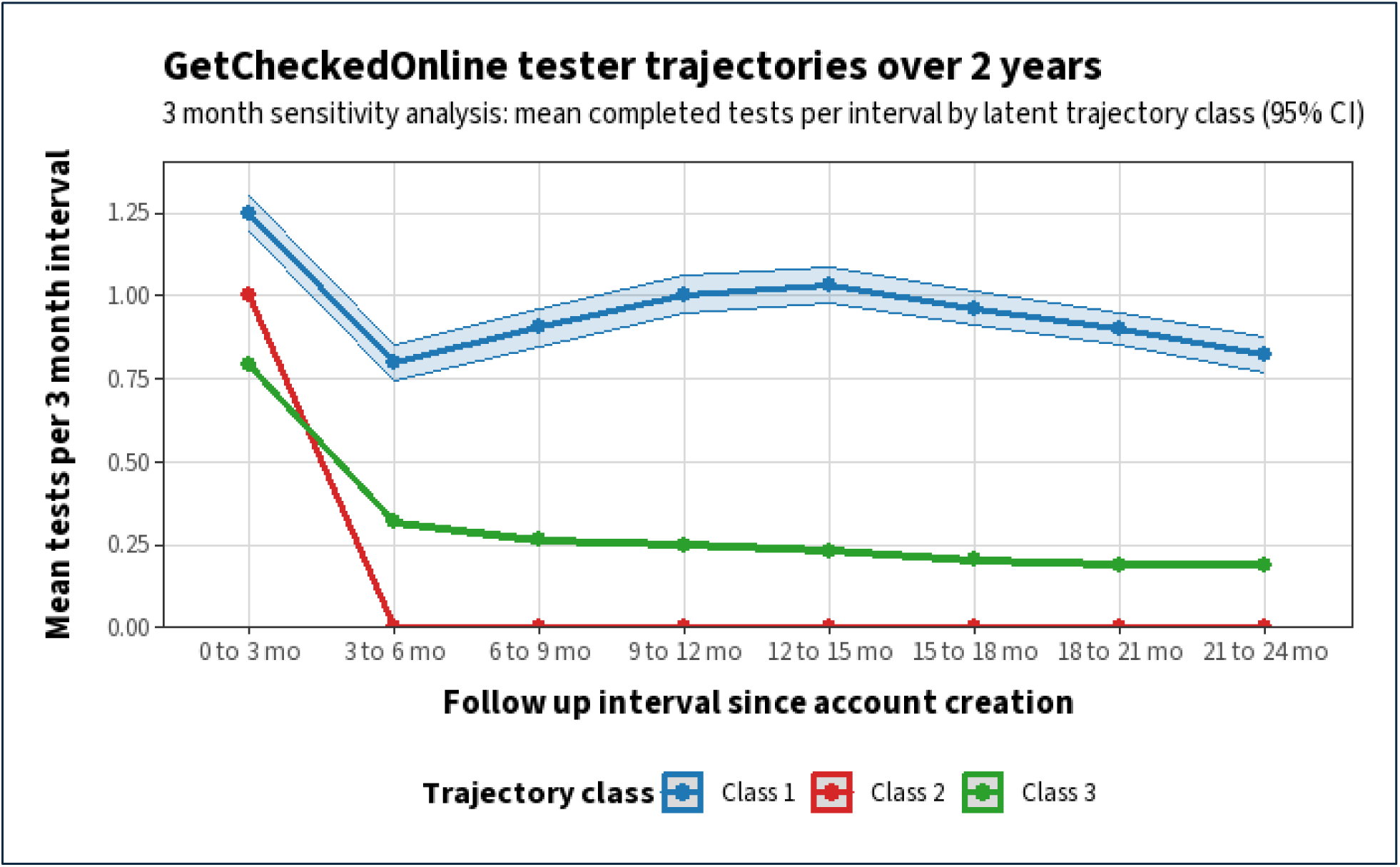
Estimated testing trajectories using 3-month time intervals.

In a sensitivity analysis using 3-month follow-up intervals, three latent testing trajectories were identified among GetCheckedOnline testers (Figure S1). Class 1 comprised 913 users (4.1%) and was characterized by sustained repeat testing across follow-up. Class 2 was the largest group, including 12,052 users (53.5%), and represented early one-time testers with no subsequent testing after account creation. Class 3 included 9,577 users (42.5%) and reflected low-frequency intermittent testing, with modest test activity that declined gradually over time.

Compared with the primary analysis using 6-month intervals, the high-use repeat testing trajectory remained stable. In contrast, the lower-frequency trajectory observed in the primary analysis subdivided into early one-time testers and intermittent testers under finer temporal resolution. This suggests that the primary two-class solution captures the dominant population-level testing patterns, while shorter follow-up intervals reveal additional heterogeneity within lower-use groups without altering the overall structure of testing behavior.

#### Sensitivity analysis 2 – Group-based trajectory modeling excluding highest frequency testers

**Supplemental Figure S2:**
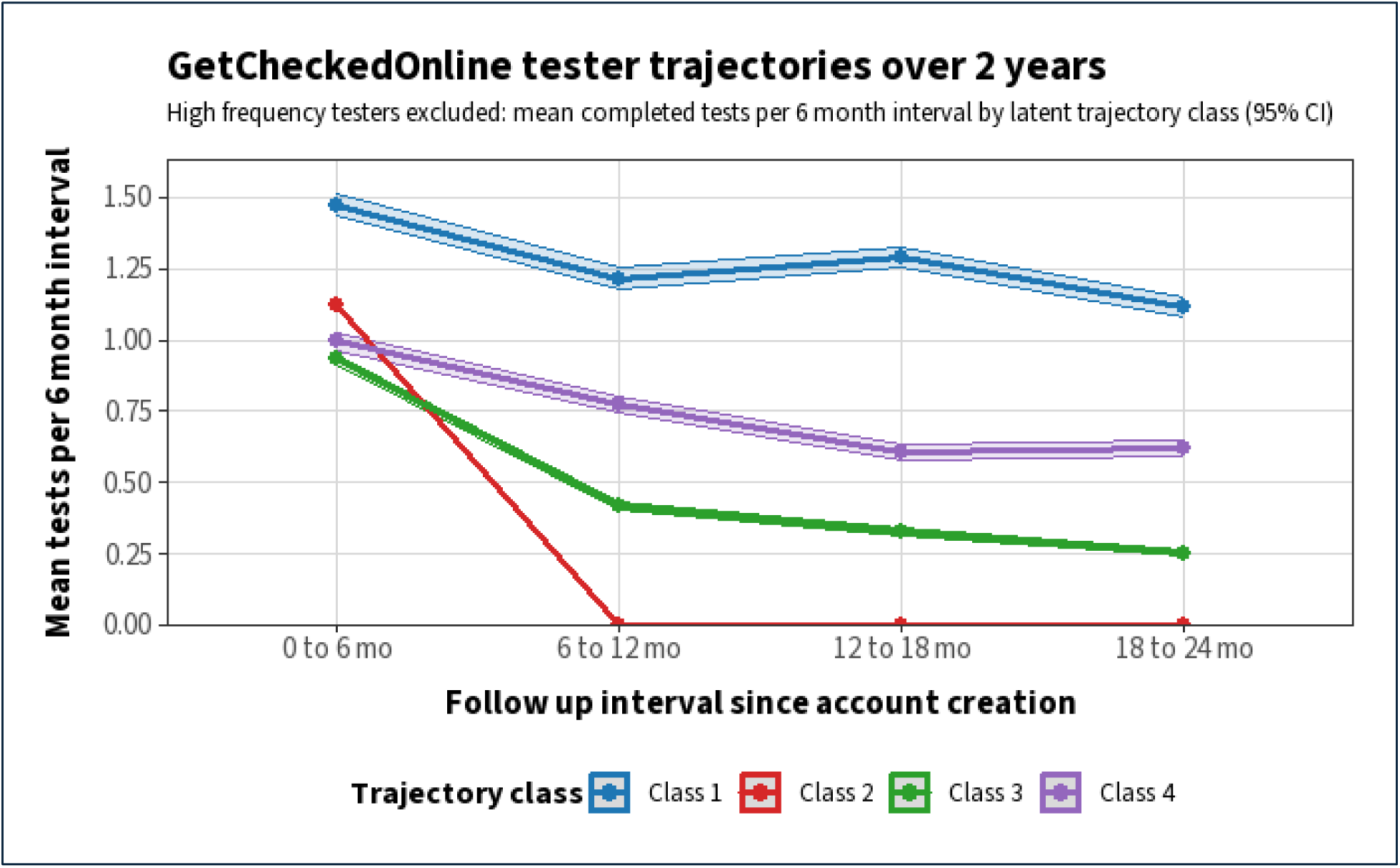
Estimated testing trajectories after excluding users above the 99th percentile of testing frequency.

To assess whether trajectory structure was driven by a small number of extreme high-frequency users, we repeated the trajectory analysis after excluding testers above the 99th percentile of completed testing episodes. In this analysis, four latent testing trajectories were identified (Supplementary Figure S2). Class 1 comprised 1,965 users (9.0%) and was characterized by sustained repeat testing across follow-up, though at lower absolute intensity than the high-use group observed in the full cohort. Class 2 was the largest group, including 12,785 users (58.8%), and represented early one-time testers with little to no subsequent testing after account creation. Class 3 included 5,115 users (23.5%) and reflected low-frequency intermittent testing with gradual decline over time. Class 4 comprised 1,900 users (8.7%) and showed moderate but declining repeat testing, intermediate between sustained and low-use trajectories.

Relative to the primary analysis, exclusion of the highest-frequency testers attenuated the prominence of the sustained high-use trajectory and revealed additional heterogeneity among repeat testers. However, the dominant population-level pattern remained characterized by early one-time testing. These findings indicate that the primary trajectory structure is not driven solely by extreme high-frequency users and that meaningful variation in testing behavior persists even after their exclusion.

#### Sensitivity Analysis 3 – Testing alternative trajectory specifications

We further assessed the robustness of trajectory classification by estimating models with alternative functional forms (linear, quadratic, cubic). Model fit statistics and selected class solutions are summarized in Table S X.

**Supplementary Table S2:** Model fit and class solutions across alternative trajectory specifications.

| **Trajectory specification** | **Selected number of classes (k)** | **BIC** | **AIC** | **Class sizes (n)** |
| --- | --- | --- | --- | --- |
| Linear | 2 | 137,178.90 | 137,131.90 | Class 1: 7,066; Class 2: 15,476 |
| Quadratic | 4 | 134,996.50 | 134,930.60 | Class 1: 20,359; Class 2: 2,183 |
| Cubic | 4 | 134,667.40 | 134,582.70 | Class 1: 20,507; Class 2: 2,035 |

Although quadratic and cubic specifications selected four statistical components based on information criteria, inspection of class-specific trajectories and posterior class membership distributions showed that these higher-order solutions consistently collapsed into two substantively distinct testing patterns corresponding to higher-frequency repeat testers and lower-frequency testers. The additional components reflected minor subdivisions within dominant trajectories rather than qualitatively distinct behavioral profiles. Accordingly, the two-class solution was retained as the primary analytic framework due to its interpretability, parsimony, and consistency across model specifications.

**Supplementary Table S3:** Model fit and variance explained for stepwise multilevel models predicting Class 1 membership (All users)

| **Metric** | **Null** | **Model 1:**<br>**Individual**<br>**factors** | **Model 2: +**<br>**System**<br>**factors** | **Model 3: +**<br>**Structural**<br>**context** |
| --- | --- | --- | --- | --- |
| N | 34,217 | 34,217 | 34,217 | 34,217 |
| AIC | 25,170 | 24,476 | 24,482 | 24,476 |
| R<sup>2</sup> marginal (Nakagawa) | 0 | 0.086 | 0.092 | NA |
| R<sup>2</sup> conditional (Nakagawa) | 0.042 | 0.117 | 0.109 | NA |
| Tjur R<sup>2</sup> | 0.005 | 0.024 | 0.024 | 0.024 |
| ICC | 0.042 | 0.034 | 0.029 | NA |
| Model type | GLMM | GLMM | GLMM | GLM |

R<sup>2</sup> marginal represents variance explained by fixed effects; R<sup>2</sup> conditional represents variance explained by fixed and random effects combined. ICC indicates the proportion of total variance attributable to differences between LHAs. The final model was estimated as a fixed-effects logistic regression due to negligible between-area variance. GLMM = generalized linear mixed model (including random effects); GLM = generalized linear model (fixed effects only).

**Supplementary Table S4:** Model fit statistics for sequential regression models predicting higher-intensity testing trajectory membership (Testers Only).

| **Model** | **Model type** | **Marginal**<br>**R<sup>2</sup>** | **Conditional**<br>**R<sup>2</sup>** | **Tjur's R<sup>2</sup>** |
| --- | --- | --- | --- | --- |
| Demographic + risk | Mixed-effects logistic regression | 0.45 | 0.45 | — |
| Demographic + risk + structural | Fixed-effects logistic regression | — | — | 0.23 |
| Demographic + risk + structural + community | Fixed-effects logistic regression | — | — | 0.24 |

Notes: Marginal and conditional R<sup>2</sup> are reported for mixed-effects models only, representing variance explained by fixed effects alone and by fixed plus random effects, respectively. For fixed-effects logistic regression models, Tjur's R<sup>2</sup> is reported as an interpretable measure of explained variance. Models including structural and community-level covariates were refit as fixed-effects models due to singular random-effects variance estimates.

